## Supplementary materials for "Oxytocin administration enhances pleasantness and neural responses to gentle stroking but not moderate pressure social touch by increasing peripheral concentrations" (from another version of this preprint: version 1, 2023-01-05)

### Supplemental information

#### Chen et al.,

### Methods

#### Questionnaires

Before the treatment administration, all participants were instructed to complete Chinese versions of validated questionnaires on individual personality traits, mood, and attitude towards interpersonal touch and sensitivity to reward. Personality trait measures included the Beck Depression Inventory II (BDI) (Beck et al., 1996), the State-Trait Anxiety Inventory (STAI) (Spielberger et al., 1983), the Cheek and Buss Shyness Scale (CBSS) (Cheek and Buss, 1981), the Interpersonal Reactivity Index (C-IRI) (Siu and Shek, 2005), the Childhood Trauma Questionnaire (CTQ) (Bernstein et al., 2003), and the Adult Attachment Scale (AAS) (Collins and Read, 1990). Individual attitudes and sensitivity to touch and reward were assessed using the Social Touch Questionnaire (STQ) (Wilhelm et al., 2001), the Sensitivity to Punishment and Sensitivity to Reward Questionnaire (SPSRQ) (Torrubia et al., 2001), and individual levels of sensory over-responsivity (also referred to as defensive responses) towards tactile stimuli was measured by the Sensory Over-Responsivity (SensOR) Scales (Schoen et al., 2008). To provide a measure of autistic traits to investigate associations with behavioral and neural results subjects completed the Autism Spectrum Quotient (ASQ) (Baron-Cohen et al., 2001). Additionally, to control for potential confounding effects of treatment on mood, participants completed the Positive and Negative Affect Schedule (PANAS) (Watson et al., 1988) immediately before and 30 min after the treatment and after each massage/touch session (details see **Figure 1A**).

#### Blood samples and OT assay

Blood samples (6 ml) were collected in EDTA tubes before and 30 min after treatments, immediately after each stimulation session using venipuncture. Oxytocin concentrations were measured using a commercial ELISA (ENZO, USA, kit no: ADI-901-153). Blood samples were analyzed in duplicate and a standard prior extraction step was performed following the recommended protocol from the manufacturers. Spiked samples (with 100 pg/ml OT added) were included with every assay to calculate extraction efficiency which was 96%. The extraction step incorporated a 4-fold concentration of samples using a vacuum concentrator (Concentrator plus, Eppendorf, Germany) resulting in a detection sensitivity of 2 pg/ml. All samples had detectable concentrations. The inter- and intra-assay coefficients of variation were 14 and 11% respectively. The manufacturer's reported cross-reactivity of the antibody for other neuropeptides, such as vasopressin and vasotocin, is < 0.01% (for detailed OT assay methods, see Li et al., 2019).

**Experimental task**

The task started approximately 35-40 min after the treatment administration. A professional masseur blinded to the research aim was trained by the experimenter to administer the different social touch stimuli to the calf of each leg as consistently as possible. For gentle touch the masseur applied only a gentle stroking touch at the optimum velocity for activating CT-fibers (5cms/s) and at which subjects perceive this form of touch as most pleasant (Löken et al., 2009). For the massage stimulation condition the masseur applied a medium pressure massage at the same velocity which while it should also activate CT-fibers to some extent was designed to primarily include a medium pressure component targeting non-C-type fibers (Field, 2010, 2019). The gentle touch and medium pressure massage were delivered on both legs simultaneously to control for possible preferences for left or right and more importantly to avoid unilateral brain activation. During the experiment, the masseur could simultaneously see a visual cue indicating the type of stimulation on a personal monitor and was instructed to vary the exact start and end point of each stimulation on the calf randomly by a few millimeters in order to minimize receptor fatigue (cf. Cascio et al., 2012). To reduce the possibility that subjects might be uncomfortable with receiving gentle caress-like touch from a stranger, subjects were informed that both types of touch stimuli they would receive were forms of professional massage using different amounts of pressure and that they just needed to be relaxed and concentrate on how the administered 'massage' made them feel. They were further informed that either a masseur or a masseuse would be randomly assigned by the experimenter to deliver the 'massage', although in fact, they were always given by a same masseur.

### Results

#### Treatment effects on mood

To determine treatment effects on individual mood, repeated-measures ANOVAs with time (pre-treatment vs. post-treatment vs. post-medium pressure massage vs. post-gentle stroking touch) as within-subject factor and treatment (intranasal OT vs. oral OT vs. PLC) as between-subject factor were employed separately for the positive and negative mood scores. For the positive mood, results showed a significant main effect of time ($F(3, 504) = 11.12$, $p < 0.001$), with significant decrease of the positive mood from pre-treatment to post- pressure massage ($p = 0.003$) and post-gentle stroking touch ($p < 0.001$). For the negative mood, there was also a significant main effect of time ($F(3, 504) = 27.10$, $p < 0.001$), with significant decrease of negative mood from post-treatment to post- medium pressure massage ($p < 0.001$) and post-gentle stroking touch ($p < 0.001$). For both sub-scale scores, there were no significant main effects of treatment and interactions (*p*s > 0.13).

#### Physiological results

Repeated-measures ANOVA with treatment as between-subject factor and stimulation condition as within-subject factor was conducted for the HRV measures (N = 168, data of 4 subjects were not recorded correctly). Results of the autonomic, parasympathetic functioning indexed by the DFAα1 revealed a significant main effect of condition ($F(1, 164) = 16.02$, $p < 0.001$, $\eta_p^2 = 0.09$) with higher DFAα1 (0.97 ± 0.27) following the applied gentle stroking touch in comparison with the medium pressure massage (0.89 ± 0.26) (**Table 1**). No significant main effects of treatment ($F(2, 164) = 0.33$, $p = 0.72$) and interactions ($F(2, 164) = 0.11$, $p = 0.89$) were found.

For the SCR data analysis (N = 165), 6 subjects were excluded from the analysis due to the low-quality data), there was no significant difference in SCR amplitude between different treatment groups ($F(2, 162) = 0.95$, $p = 0.39$) and the treatment x condition interaction was not significant ($F(6, 162) = 0.43$, $p = 0.86$, see **Table 1**). However, results showed a significant main effect of condition ($F(3, 162) = 85.01$, $p < 0.001$, $\eta_p^2 = 0.34$) with a significant increase of SCR amplitude following both the gentle stroking touch and the medium pressure massage compared to rest periods (*p*s < 0.001) and higher SCR amplitude induced by the medium pressure massage than the gentle stroking touch ($p < 0.001$).

**Effects of perceived gender on behavioral pleasantness rating scores**

Based on the debrief after completing the task, a total of 94 subjects thought the received massage was administered by a male masseur (N = 77 thought by a female) but they did not know the massage was in fact always from the same male masseur.

To explore whether the perceived gender of the masseur would influence the pleasantness rating scores, repeated-measures ANOVA with the perceived gender as the between-subject factor and the condition as the within-subject factor found no significant main effect of the perceived gender ($F(1, 169) = 0.60$, $p = 0.44$, $\eta_p^2 = 0.004$) and interactions ($F(1, 169) = 0.22$, $p = 0.64$, $\eta_p^2 = 0.001$). Additionally, OT administration did not change the individual perception of the gender of the person applying the stimulation ($\chi^2 = 1.63$, $p = 0.44$). And the effects of OT on increased pleasantness ratings of the gentle stroking touch was similar no matter whether participants thought they had received the touch from a male or female since results showed no significant main effect ($F(1, 165) = 0.83$, $p = 0.36$, $\eta_p^2 = 0.005$) and interaction effects of the perceived gender ($p$s > 0.48) apart from the still significant main effect of treatment ($F(2, 165) = 3.93$, $p = 0.02$, $\eta_p^2 = 0.046$) and the significant treatment x condition interactions ($F(2, 165) = 3.36$, $p = 0.037$, $\eta_p^2 = 0.04$).

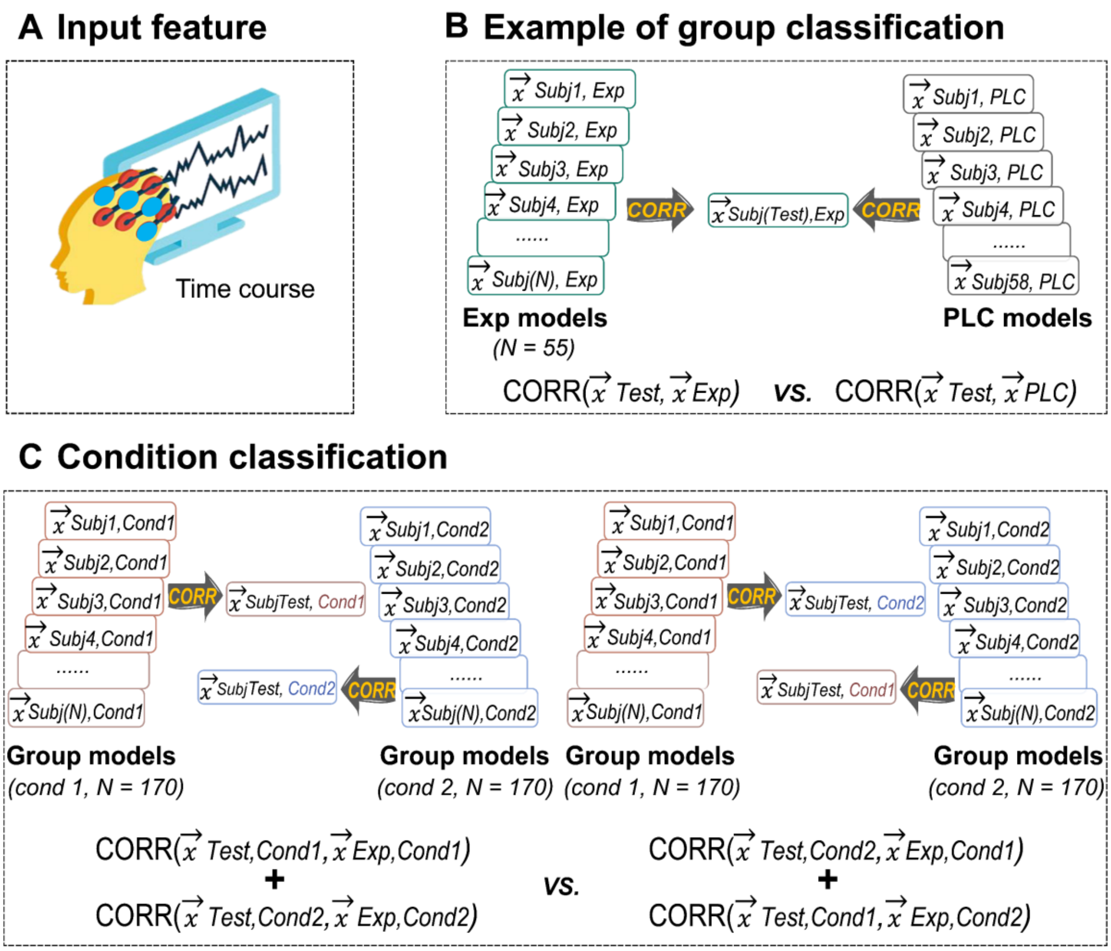


**Figure S1.** Illustration of the multivariate analysis method. (**A**) Individual mean time course (5-35s following the gentle touch stimulation onset) was used to discriminate (**B**) treatment groups and (**C**) experimental conditions. **(B)** A specific example of group discrimination between intranasal OT and PLC groups. Each subject (test subject, leave-one-out method) in intranasal OT or PLC groups was classified once at a time, by comparing the correlations of their individual activation patterns with averaged activation patterns of the same treatment group (Exp group models) versus with averaged patterns of another group (i.e. if the test subject was from intranasal group and CORR ($\vec{x}\ Test, \vec{x}\ Exp$) > CORR ($\vec{x}\ Test, \vec{x}\ PLC$), then the accuracy for this subject was 1). **(C)** Discrimination accuracy between two stimulation conditions was 0.73, $p < 0.001$, permutations = 10000.

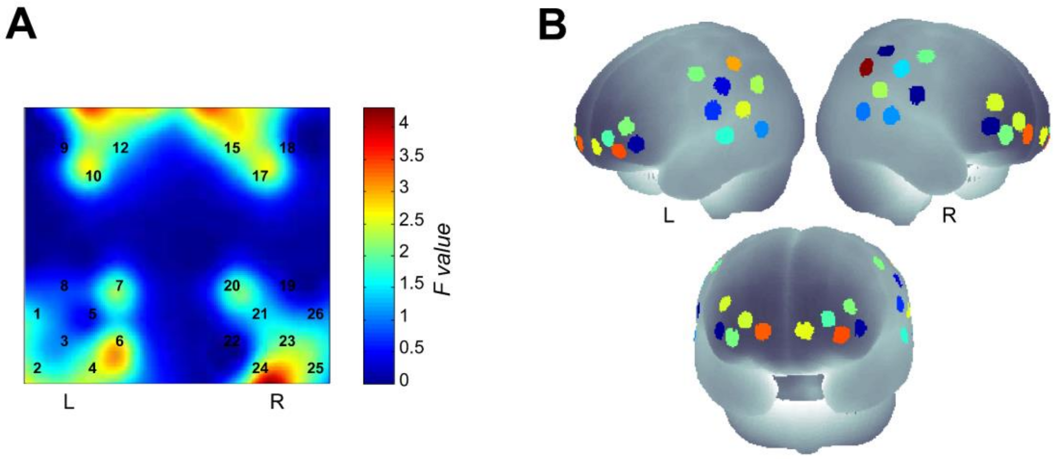


**Figure S2.** (**A**) 2D and (**B**) 3D group level activation map showing treatment x condition interaction effects at each channel. All colors represent F-statistic values of channel-by-channel group-level repeated-measures ANOVAs.
